# Population-Specific Polygenic Risk Scores Developed for the Han Chinese

**DOI:** 10.1101/2024.10.14.24315279

**Authors:** Hung-Hsin Chen, Chien-Hsiun Chen, Ming-Chih Hou, Yun-Ching Fu, Ling-Hui Li, Che-Yu Chou, Erh-Chan Yeh, Ming-Fang Tsai, Chun-houh Chen, Hsin-Chou Yang, Yen-Tsung Huang, Yi-Min Liu, Chun-yu Wei, Jen-Ping Su, Wan-Jia Lin, Elin H.F. Wang, Chi-Lu Chiang, Jeng-Kai Jiang, I-Hui Lee, Kung-Hao Liang, Wei-Sheng Chen, Hung-Cheng Tsai, Shih-Yao Lin, Fu-Pang Chang, Hsiang-Ling Ho, Yi-Chen Yeh, Wei-Cheng Tseng, Ming-Hwai Lin, Hsiao-Ting Chang, Ling-Ming Tseng, Wen-Yih Liang, Paul Chih-Hsueh Chen, Yu-Cheng Hsieh, Yi-Ming Chen, Tzu-Hung Hsiao, Ching-Heng Lin, Yen-Ju Chen, I-Chieh Chen, Chien-Lin Mao, Shu-Jung Chang, Yen-Lin Chang, Yi-Ju Liao, Chih-Hung Lai, Wei-Ju Lee, Hsin Tung, Ting-Ting Yen, Hsin-Chien Yen, Ming-Yao Chen, Ying-Chin Lin, Yung-Ta Kao, Bi-Zhen Kao, Jing-Er Lee, Chi-Li Chung, Ju-Chi Liu, Paul Chan, Chang-Hsien Lin, Chia-Hsin Chen, I-Chen Wu, Lung-Chang Lin, Jiunn-Wei Wang, Shen-liang Shih, Sun-Wung Hsieh, Chih-Hsing Hung, Wei-Ming Li, Chih-Jen Yang, Cheng-Shin Yang, Ru-Hui Weng, Yu-Chi Chen, Chun-Ping Chang, Tai-Hsun Wu, Yu-Chang Lin, Yi-Jing Sheen, Shi-Heng Wang, Sye-Pu Chen, Timothy Raben, Erik Widen, Stephen Hsu, Feng-Jen Hsieh, Dong-Ru Ho, Yu-Huei Huang, Chung-Han Yang, Yu-Shu Huang, Yen-Fu Chen, Hsien-Ming Wu, Ping-Han Tsai, Kuan-Gen Huang, Chih-Yen Chien, Yi-Lwun Ho, Ming-Shiang Wu, Jia-Horng Kao, Yen-Bin Liu, Jyh-Ming Jimmy Juang, Mao-Hsin Lin, Yen-Hung Lin, Ji-Yuh Lee, Hsueh-Ju Lu, Chieh-Hua Lu, An-Chieh Feng, Jhih-Syuan Liu, Chien-Ping Chiang, Nain-Feng Chu, Jung-Chun Lin, Yi-Wei Yeh, En Meng, Chih-Yang Huang, Chi-Cheng Li, Tso-Fu Wang, Kuei-Ying Su, Jia-Kang Wang, Mei-Hsiu Chen, Hua-Fen Chen, Gwo-Chin Ma, Ting-Yu Chang, Fu-Tien Chiang, Hsing-Jung Chang, Kuo-Jang Kao, Chen-Fang Hung, Ching-Yao Tsai, Po-Yueh Chen, Kochung Tsui, Yuan-Tsong Chen, Pui-Yan Kwok, Wayne Huey-Herng Sheu, Shun-Fa Yang, Jyh-Ming Liou, Jaw-Yuan Wang, Jeng-Fong Chiou, Jer-Yuarn Wu, Cathy S.-J. Fann

## Abstract

Predicting complex disease risks based on individual genomic profiles is an advancing field in human genetics^1,2^. However, most genetic studies have focused on European populations, creating a global imbalance in precision medicine and underscoring the need for genomic research in non-European groups^3,4^. The Taiwan Precision Medicine Initiative (TPMI) recruited over half a million Taiwanese residents, providing a large dataset of genetic profiles and electronic medical record data for the Han Chinese. Using extensive phenotypic data, we conducted comprehensive genomic analyses of Han Chinese across the medical phenome. These analyses identified population-specific genetic risk variants and novel findings for various complex traits. We developed polygenic risk scores, demonstrating strong predictive performance for conditions such as cardiometabolic diseases, autoimmune disorders, cancers, and infectious diseases. We observed consistent findings in an independent dataset, Taiwan Biobank, and among East Asians in the UK Biobank and the All of Us Project. The identified genetic risks accounted for up to 10.3% of the overall health variation in the TPMI cohort. Our approach of characterizing the phenome-wide genomic landscape, developing population-specific risk prediction models, assessing their performance, and identifying the genetic impact on health, serves as a model for similar studies in other diverse study populations.

## Introduction

A major promise of modern genetics is the ability to predict complex disease risk based on a person’s genetic profile. If successful, health management strategies can be developed to mitigate the risk (disease prevention) and to optimize care (early diagnosis and effective treatment). Large-scale studies by the UK Biobank (UKB) and Electronic Medical Records and Genomics (eMERGE) Network show that risk prediction based on genetics holds promise and several countries are exploring ways to implement risk-based management in clinical practice^1,2^. Using polygenic risk scores (PRS) to predict disease risk and identify individuals at high risk is an emerging “precision medicine” approach to leverage genetic findings in clinical practice. However, a significant limitation is that current PRS models are predominantly based on genome-wide association studies (GWAS) with participants of European ancestries (EUR)^4,5^, often leading to reduced predictive performance in groups of other anestry^6,7^. To fully realize the potential of precision medicine for diverse global populations, population specific phenome-wide genomic discovery must be perfomed at scale and clinically applicable polygenic risk models must be optimized within and across populations. To fill this major research gap in an population of East Asian ancestry (EAS), we characterized the complex genetic architecture of the population of Han Chinese ancestry phenome-wide, developed population-specific PRS, and assessed the external validity of the models across populations with varying degrees of genetic similarity.

Populations of EAS ancestry represent nearly a quarter of the global population, but they account for only 3.95% of the participants in previous GWAS^3^. Although several biobanks have been built to recruit subjects from East Asia, they have moderate sample size (72K-212K) and many focus on specific conditions^8–12^. In contrast, biobanks with predominantly European ancestry participants^13–16^, have significantly larger sample sizes (224K-635K) and access to more comprehensive clinical data. The moderate sample size and limited phenotypes in existing EAS biobanks hamper discovery of unique genetic effects and preclude the development of robust and clinically useful PRS models for EAS.

We assembled a large non-EUR cohort, the Taiwan Precision Medicine Initiative (TPMI), and genotyped over half a million participants across sixteen medical centers in Taiwan from 2019 to 2023. All the participants, who are overwhelmingly of Han Chinese ancestry, contributed DNA samples for genetic profiling with a custom-designed genotyping array and consented to provide their longitudinal electronic medical records (EMR) from 5 years prior to enrollment and into the future. The EMR dataset includes rich and accurate health-related phenotypes, including medical diagnoses and biochemical examinations^17^. Here, we present the results of comprehensive genomic analyses with extensive genetic and medical data derived from the TPMI cohort, including phenome-wide GWAS and PRS model development. We identified numerous population-specific risk variants/genes, observed evidence of genetic pleiotropy, and pinpointed clusters of traits that shared similar genetic etiology. Then, we developed and validated PRS prediction models for numerous conditions against external datasets including those from the Taiwan Biobank (TWB), the UKB, and the All of Us Project. Our results reveal the benefits of leveraging a large cohort from understudied population to identify unique genetic underpinnings of the human phenome, interpret causal effects via fine mapping and colocalization, and improve the performance of population-specific PRS models, which together, better illuminate the clinical implications of genetic risk.

## Results

### Diseases and Quantitative Traits in TPMI

We performed comprehensive genomic analyses, including GWAS, heritability estimation, and PRS model building and evaluation, across a wide range of diseases and quantitative traits using 463,447 genetically inferred Han Chinese from TPMI. We examined 695 dichotomized phenotypes (phecode, case n >2,000) and 24 quantitative traits (sample size >100,000), spanning numerous disease categories (defined by phecode groupings^18,19^), such as neoplasms, metabolic disorders, circulatory conditions, autoimmune diseases, and more (Fig. 1). The phecodes, derived from International Classification of Diseases (ICD) codes^18,19^, alongside quantitative traits such as blood pressure, BMI, liver enzymes, and lipid levels, provide a robust dataset for exploring genetic contributions to human health (Table S1 and S2). The log-transformed case proportion identified from EMR showed a moderate but significant correlation with the log-transformed 5-year disease prevalence from National Health Insurance Research Database (NHIRD) in Taiwan^20^ (r = 0.656, p-value = 2.69×10^−84^) (Fig. 1A and Extended Data Fig. 1), suggesting that the TPMI’s hospital-based design may not fully capture mild and common illness, which are primarily observed in local and primary care clinics. Fig. 1B displays the sample sizes for 24 quantitative traits in the TPMI and highlights sample size variation across traits, a key measure impacting the power and precision of association analyses within the cohort.

**Fig. 1.**
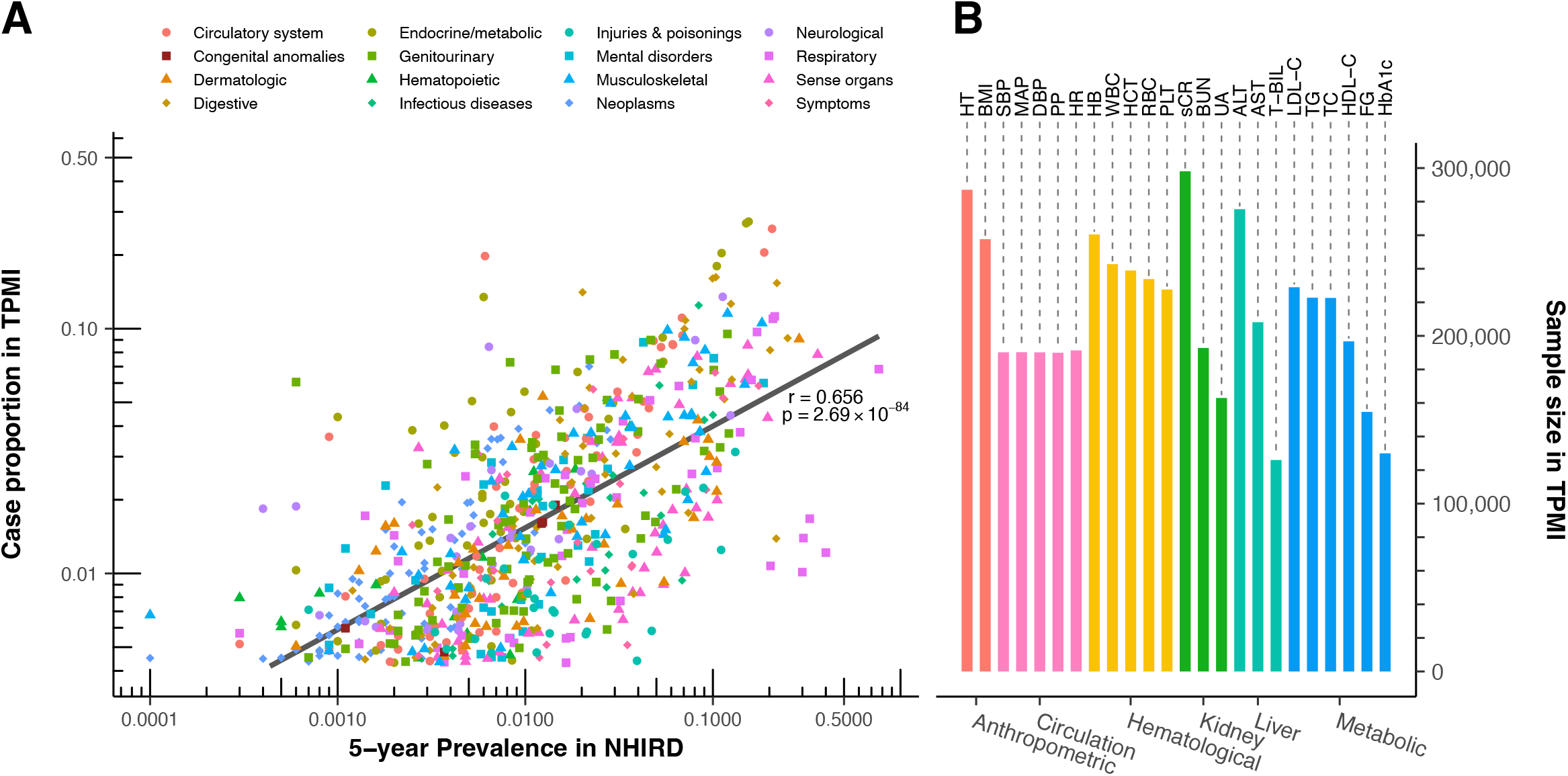
Scatter plot of the case proportion for dichotomized phenotypes and barchart of sample size for quantitative traits in TPMI dataset. (A) The case proportion in TPMI is compared to the 5-year prevalence in the National Health Insurance Research Database (NHIRD) for dichotomized phenotypes (phecodes). Each dot represents a specific phecode, with the x-axis showing the prevalence in NHIRD and the y-axis showing the case proportion in TPMI. The correlation coefficient and significance were calculated using the two-sided Pearson correlation test. (B) Scatter plot shows the sample sizes for quantitative traits in the TPMI cohort. Each point represents a trait, with the x-axis indicating the different category of quantitative traits and the y-axis representing the corresponding sample sizes.

### GWAS, Fine-Mapping, and Novel Results

Our GWAS identified at least one significant locus (p-value < 5×10^−8^) for 265 phecodes of the 695 tested and all 24 quantitative traits. Highlighting the robustness of the TPMI data, we observed a high replication rate of reported disease loci from EAS GWAS on GWAS catalog (actual/expected ratio (AER) = 78.17%, considering the statistical power with the published tool, PGRM^21^), particularly for endocrine and metabolic/hematopoietic diseases (AER = 88.68% and 84.62%, respectively; Extended Data Fig. 2 and Table S3). Lower replication rates for respiratory (AER = 23.53%) may reflect limited case numbers, untyped genetic variants, such as rare variants, copy number variation, and structural variants, or recruitment bias such as age distribution. We applied the sum of single effects model for fine-mapping to identify the independent variant-trait associations, and reported the genetic variant with highest posterior inclusion probability of identified credible sets as well as the single lead variant for the failed fine-mapping regions and major histocompatibility complex region (MHC region, chr6:25M-33M). Our analyses revealed a total number of 2,656 fine-mapping identified independent association signals, including 1,309 from phecodes GWAS and 1,347 from quantitative traits. Notably, 95 novel associations, defined as having no previously reported results within 1Mb in the NHGRI-EBI GWAS Catalog^21^ of relevant GWAS, were identified across 50 phecodes and 7 quantitative traits. In addition, we identified 217 new hits from previously reported regions, defined as having r^2^<0.1 with any variant observed in the NHGRI-EBI GWAS Catalog within 1 MB for the same phenotype (Table S4 and S5). After applying multiple test correction, 1,502 fine-mapped associations passed Bonferroni-adjusted threshold (5×10^−8^/(695+24) = 6.95×10^−11^), as well as 21 novel variants and 115 new hits.

Of the 95 novel genetic associations, 30 variants are rare (minor allele frequency [MAF]<0.05) in populations with other ancestry (with African ancestry, Admixed American ancestry, South Asian ancestry, and European ancestry (non-Finnish) in gnomAD), and 33 variants less than 0.01 in the EUR, the most extensively studied population, which explains why they were not reported in previous GWAS. For example, the SNP rs17089782, a missense variant in the *PIBF1* (p.R405Q) on chromosome 13 is significantly associated with thyroid cancer (p = 2.8×10^−9^) in the TPMI cohort. This SNP has a MAF of 5.65% in TPMI but 0.01% in EUR, which may explain why this association was only detectable in TPMI. However, *PIBF1* is essential for immune regulation, especially during pregnancy, and is relevant to autoimmune diseases and cancer^22^. Another novel variant identified in our quantitative analysis of BMI (rs761018157 p-value = 4.8×10^−9^, MAF in TPMI = 4.34%, MAF in EUR<0.01%,) maps to *PHOX2B*. This gene, highly expressed in the nervous system, had previously been linked to obesity hypoventilation syndrome in a small study (n = 30)^23^ and associated with bone mineral density^24^. In addition, when we compare the effect size in TPMI and UKB for the rest of the novel findings, 25 exhibit a significant different effect size (p-value<0.05). For instance, a TPMI-identified platelet count-associated variant, rs12955741, located in the intergenic region between *TGIF1* and *DLGAP1* exhibits a different effect size compared to that in UKB (β_TPMI_ = 0.044, β_UKB_ = −0.005, p-value = 1.7×10^−9^). Moreover, the high hepatitis B virus (HBV) carrier rate in Taiwan^25^ contrasts sharply with its rarity in European cohorts, enabling TPMI to identify novel loci associated with viral hepatitis B (case number in TPMI = 23,618 vs. UKB = 132). Among the 26 independent loci identified in our analysis of hepatitis B, 19 fine-mapped loci are novel (Extended Data Fig. 3). Notably, 18 of these 19 loci were found to be associated with liver function or diseases (Table S5). These novel associations highlight the uniqueness of certain disease loci in the TPMI cohort, presenting opportunities for developing population-specific therapeutic interventions and advancing precision medicine.

All identified independent associations are summarized in Fig. 2. The identification of the MHC region as a significant hotspot on chromosome 6 underscores its extensive involvement in immune-related diseases across multiple categories. Similarly, the short arm of chromosome 11 (*INS*-*KCNQ1* region) also affect various traits, including metabolic, endocrine, and genitourinary diseases. These hotspots of trait-relevant variants implied the shared genetic mechanism among diseases and potential of pleiotropic effects.

**Fig. 2.**
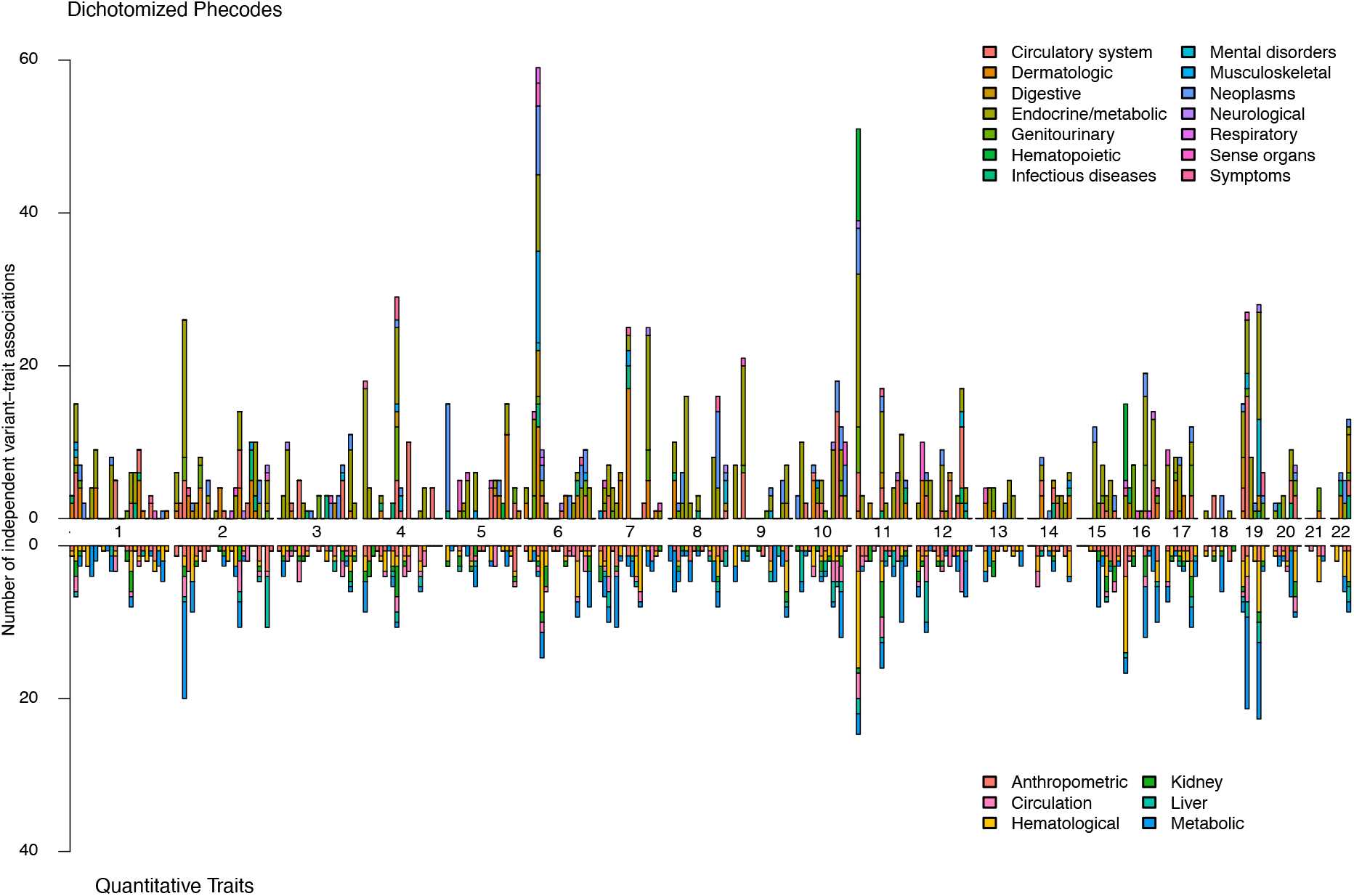
Pheno-wide independent variant-trait associations. Vertical bars show the accumulated number of independent variant-trait associations for dichotomized phecodes (top panel) and quantitative traits (bottom panel). Each category of diseases and traits is represented by a corresponding color. The X-axis is chromosome number, and the Y-axis represents the accumulated number of associations, highlighting the uneven distribution of trait-associated variants across phenotypes.

### Heritability and Colocalization

Linkage disequilibrium score regression analysis (LDSC)^26^ showed strong liability-scaled SNP-heritability for conditions such as alcoholism (h^2^ = 0.213), retention of urine (h^2^ = 0.163), and open-angle glaucoma (h^2^ = 0.160). Among quantitative traits, body height (h^2^ = 0.323), BMI (h^2^ = 0.218), and high-density lipoprotein cholesterol (h^2^ = 0.191) exhibited the highest heritability estimates (Table S6 and Supplmentary Information), highlighting the significant role of genetics in these traits. These results have far-reaching implications for precision medicine, as higher heritability signals suggest the potential for more accurate genetic risk prediction models that could improve personalized disease risk assessments.

We then partitioned the heritability at the gene level and identified 329 unique genes contributing significantly to phenotypic variation (h^2^ >0.1% and z-score >1.64). Of these, 45 affected more than one phecode and/or quantitative category, including key genes such as *APOE, APOC1, TOMM40, ABCG2*, and *KCNQ1* (Fig. 3 and Table S7). We also conducted a colocalization analysis to elucidate the potential molecular function of identified GWAS signals with three QTL datasets, including GTEx^27^, MAGE^28^, and JCTF^29^ (Fig. 3 and Table S8). Our results identified 391 unique genes that potentially mediate the outcome through their expression level (posterior probability > 0.9), including *GBAP1* which colocalized with five different traits (uric acid, serum creatinine, hematocrit, hypertension, and gout). Among the colocalized genes, 75 of them can be identified only in the multi-ancestry lymphoblastoid cell lines eQTL, MAGE (20 genes), and/or Japanese whole blood eQTL, JCTF (59 genes). Our findings demonstrate the effect of these genes (such as *APOE, ABCG2*, and *KCNQ1*) on multiple traits and disorders. By elucidating shared genetic effects, these results offer opportunities to develop precision medicine approaches that address comorbidities, such as treating hyperlipidemia and reducing dementia risk through a single intervention targeting *APOE*, which influences both lipid metabolism and Alzheimer’s disease risk. This gene-level understanding emphasizes the potential to optimize therapeutic strategies by leveraging genetic pleiotropy in disease management.

**Fig. 3.**
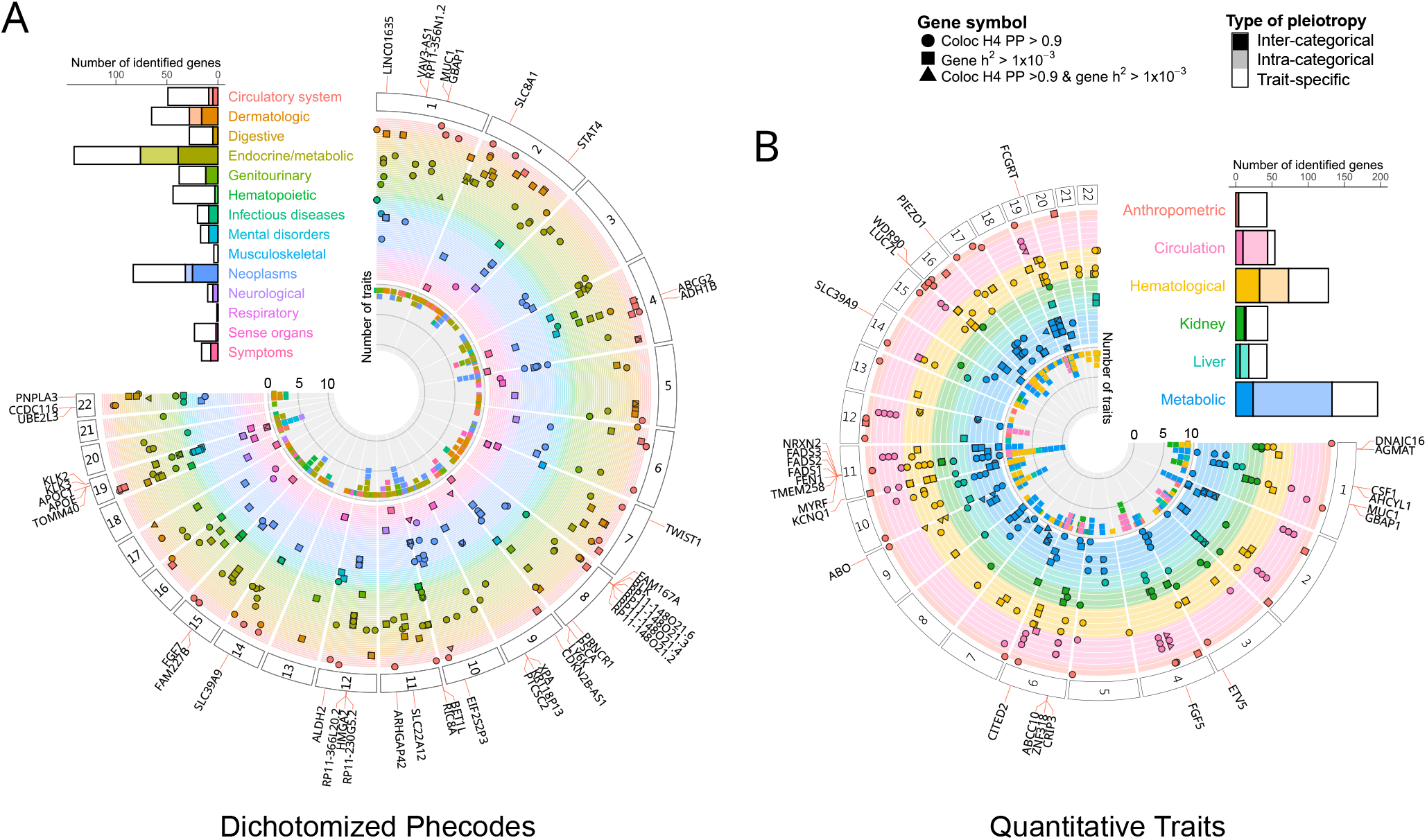
Gene-level heritability and colocalization with gene expression. Circle plot showing gene-level heritability and colocalization with gene expression for (A) dichotomized phenotypes, summarized in parent (integer) phecodes, and (B) quantitative traits. Dots represent gene-level heritability (h^2^) > 10^−3^, squares indicate colocalization posterior probability > 0.9, and triangles show both. Inner circle indicates the number associated traits for each identified gene. The bar chart shows the number of identified genes by category and grouped by type of pleiotropy. The gene-level heritability was estimated with h2gene^57^, and the colocalization analysis was performed with coloc^58^.

### Genetic Correlation and Clusters

Pairwise genetic correlation analyses revealed three major phenotype clusters: cardiometabolic traits, autoimmune and infectious diseases, and kidney-related traits (Fig. 4 and Extended Data Fig. 4). The cardiometabolic cluster, which includes type 2 diabetes, hypertension, and BMI, reinforces the interconnected phenotypic and genetic architectures of cardiovascular and metabolic diseases. The cluster of autoimmune and infectious diseases, which includes viral hepatitis B, psoriasis, and systemic lupus erythematosus, illuminates shared immune system pathways and potential gene-pathogen interaction. The kidney-related cluster involved gout, chronic kidney disease, calculus of kidney and ureter, ankylosing spondylitis, and measures of urea nitrogen, creatinine, and uric acid. The shared genetic architecture provides opportunities to leverage the genetic risk of correlated traits while developing the PRS model.

**Fig. 4.**
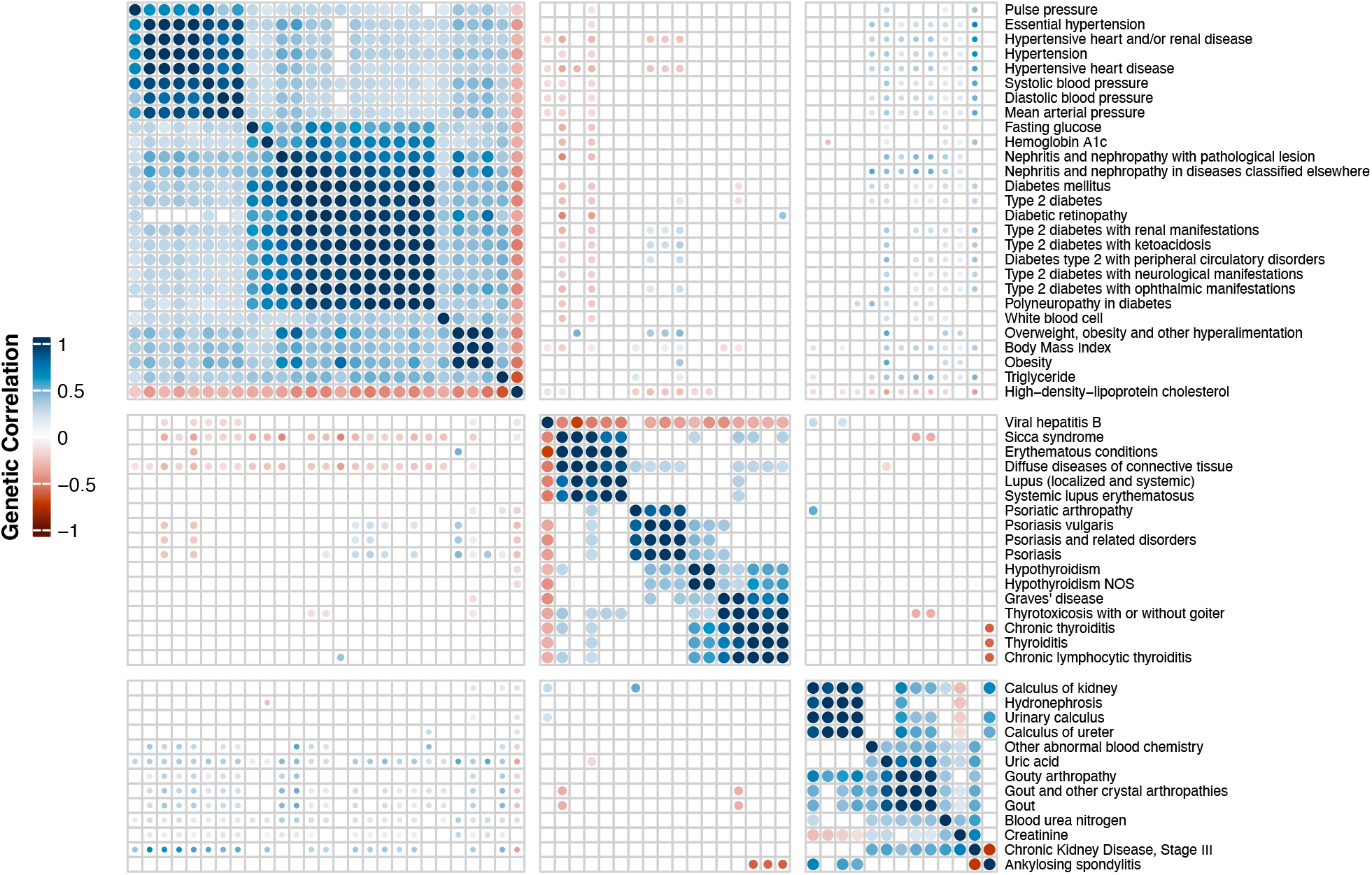
Genetic correlation among three identified trait clusters. Heatmap displays genetic correlations between trait clusters: cardiometabolic, autoimmune/infectious diseases, and kidney-related traits. Genetic correlation was estimated using LDSC, with colors representing the correlation coefficients between traits.

### Cross-Population Comparison

Cross-population comparisons^30^ with EUR GWAS from UKB showed varying degrees of genetic correlation, with strong, statistically significant correlations for traits like cholelithiasis (ρ_ge_ > 0.999), type 2 diabetes (ρ_ge_ = 0.829), and ischemic heart disease (ρ_ge_ = 0.756), but moderate correlations for gout (ρ_ge_ = 0.616) and psoriasis (ρ_ge_ = 0.418) (Table S6). The moderate correlations suggest the differentiated genetic mechanism and disease distribution across populations (gout case n = 24,411 in TPMI and 3,179 in UKB; psoriasis case n = 4,166 in TPMI and 2,197 in UKB). Therefore, these findings demonstrate the importance of population-specific genetic studies, as differences in genetic architectures between populations can significantly affect the accuracy of PRS models.

### Polygenic Risk Score (PRS) Development

Building on these insights, we developed and validated PRS models that demonstrated strong predictive performance for a wide range of diseases. Although we used five PRS tools, including LDpred2^31^, Lassosum2^32^, PRS-CS^33^, SBayesR^34^, and MegaPRS^35^ (Tables S9-S13), we found that LDpred2 outperformed the others for most traits (Extended Fig. 5). Therefore, we took the results of LDpred2 for further comparisons. Out of the 265 PRS models for phecodes, AUC values exceeded 0.55 for 105 dichotomized phecodes with a significant p-value (p-value<0.05). Additionally, the explained variance of models for 24 quantitative traits ranged from 0.028 (aspartate aminotransferase, AST) to 0.227 (height). (Table S9 and Extended Data Fig. 6). The most predictive PRS models included highly heritable traits such as ankylosing spondylitis (AUC = 0.812±0.016), psoriasis (0.709±0.016), atrial fibrillation (0.702±0.014), prostate cancer (0.696±0.018), systemic lupus erythematosus (0.696±0.015), rheumatoid arthritis (0.646±0.011), type 2 diabetes (0.640±0.005), female breast cancer (0.611±0.010), and hypertension (0.610±0.004). Interestingly, the PRS for hepatitis B also demonstrated high genetic predictability (0.654±0.008). Since SNP-heritability (h^2^) represents the upper bound of variance that can be explained by PRS, we examined the proportion of heritability captured by our models (r^2^/h^2^). A total of 36 traits, including prostate cancer (r^2^/h^2^ = 0.054/0.070), type 2 diabetes (0.066/0.126), and HDL cholesterol (0.136/0.191), reached over 50% of their SNP-heritability, suggesting that PRS can achieve near-optimal predictive accuracy for highly heritable traits. However, for complex diseases influenced by both genetic and environmental factors, PRS performance is inherently constrained by the fraction of heritability attributable to common variants. These findings reinforce the importance of SNP-heritability as a reference point for evaluating PRS utility and highlight the need for larger, ancestrally diverse datasets to further enhance genetic prediction models (Extended Data Fig. 6).

**Fig. 5.**
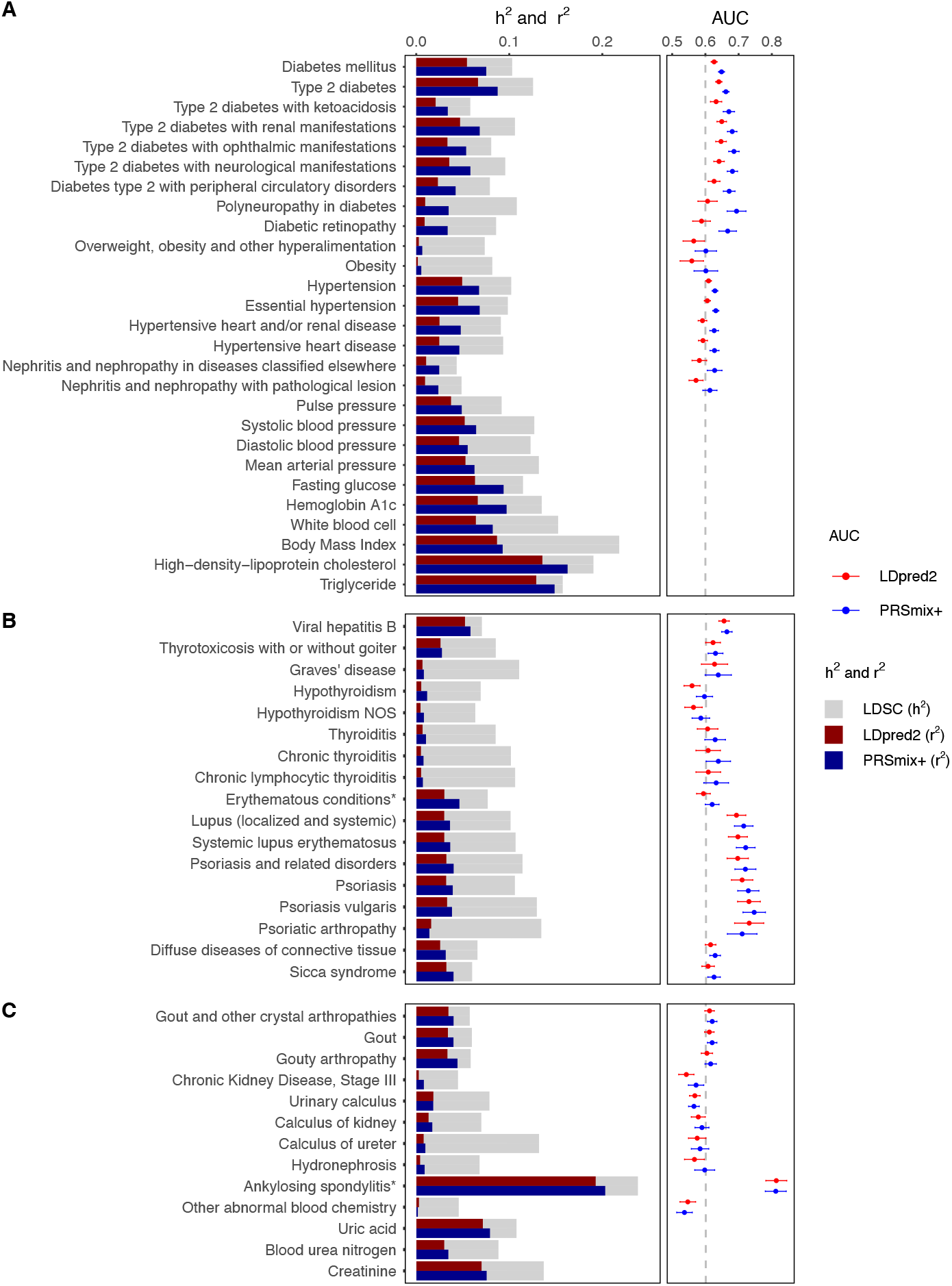
PRS performance for the three identified trait clusters. Bar plot shows SNP-heritability (h^2^) and PRS explained variance (r^2^) for (A) cardiometabolic trait cluster, (B) autoimmune trait cluster, and (C) kidney-related trait cluster. Gray bars indicate SNP-heritability (estimated from TPMI GWAS unrelated set [n=248,754] with LDSC), and the colored bar chart presents the r^2^ values, indicating the proportion of variance explained by the PRS among TPMI validation set (n=20,000) from single-trait PRS (LDpred2, red bar) or multi-trait PRS (PRSmix+, blue bar), while the dot and whisker plot showcases predictive accuracy using Area Under the receiver operating characteristic Curve (AUC) with 95% confidence interval for dichotomized traits.

Leveraging the identified clusters, we performed a multi-trait PRS training, PRSmix+^36^, for the traits within each cluster (Fig 5 and Table S14). Notably, multi-trait PRS models improved prediction accuracy for the cardiometabolic disease cluster with a 0.040 increase in AUC (from 0.608 to 0.648) and a 1.770-fold improvement in phenotypic variance explained (r^2^). The performances of autoimmune and kidney-related disease clusters were also enhanced, with averaged AUC improvements of 0.018 and 0.009, respectively (from 0.641 to 0.659, and 0.601 to 0.610 respectively), and 1.351- and 1.349-fold improvements in variance explained (r^2^). The significant enhancement of multi-trait PRS prediction (comparing r^2^ of LDpred2 and PRSmix+ with paired t-test, p = 1.07×10^−13^) highlights the potential of leveraging shared genetic architecture to enhance disease risk prediction. Fig. 5 demonstrates the performance of single and multi-trait PRS across three disease clusters, as well as the differing effectiveness of PRS in predicting genetic risk across various disease categories.

### PRS External Validation and Comparison

To evaluate the robustness and generalizability of our PRS models, we performed an external validation of the models (hypertension, type 2 diabetes, viral hepatitis B, gout, calculus of kidney from PRSmix+ and others from LDpred2) in Taiwan Biobank (TWB, genetically inferred unrelated Han Chinese, n = 88,628), UKB (self-reported EAS, n = 9,893), and All of Us (genetically inferred EAS, n = 6,895). We found that the prediction accuracy, AUC, of our models ranged from 0.548 (glaucoma) to 0.712 (prostate cancer) in TWB, 0.557 (female breast cancer) to 0.634 (hypertension) in UKB, and 0.520 (migraine) to 0.709 (gout) in All of Us (Extended Data Fig. 7). Although the TWB questionnaire did not contain specific details on hepatitis B status, we used anti-hepatitis B core total antibodies (Anti-HBc) as an indicator of infection or past infection and hepatitis B surface-antigen (HBsAg) as a marker of acute/chronic infection. Intriguingly, the AUC for the TPMI-derived model of hepatitis B were 0.674±0.003 for HBsAg and 0.530±0.002 for Anti-HBc in the TWB. These results demonstrate the high predictive value of the PRS for hepatitis B for predicting symptoms and severity of the disease.

TPMI-derived PRS models perform better than the UKB European-derived models when applied to EAS for viral hepatitis B, type 2 diabetes, hypertension, gout and migraine. (Extended Data Fig. 7) For the other traits, TPMI derived models consistently outperform the UKB ones although the confidence intervals overlap. However, the overlapping confidence intervals in UKB and All of Us may be due to their limited sample size of EAS. These results suggest that population-specific PRS models allow for more accurate risk stratification and enable personalized healthcare interventions for EAS. Additionally, we assessed the performance of TPMI-derived PRS and TPMI-included cross-population PRS across various ancestry groups, including European, African, Admixed American, and South Asian populations from the UK Biobank and All of Us cohorts (Extended Data Fig. 8). The performance varied by diseases, but consistent results were observed for female breast cancer and glaucoma across populations, and TPMI-included cross-population PRS slightly but not significantly improved in the populations other than EAS and EUR.

### Genetic Risks on Overall Health Measures

Although overall health is hard to define with a few metrics, herein, we used the count of clinical visits and duration of hospitalization to roughly describe individuals’ overall health. We found that 131 of the top performing PRS models (LDpred2 models with AUC > 0.55 and all PRSmix+ models for phecodes and all models for quantitative traits) are significantly associated with overall health indices, explaining 8.47% of the variation in clinical visit frequency (p-value = 2.69×10^−14^) and 10.29% of the variation in hospitalization duration (p-value = 5.62×10^−27^, Extended Data Table 1 and S15) in the comparison between top and bottom 5% groups after adjusting for sex, age and recruiting hospital. Among the identified clusters, the cardiometabolic disease cluster contributed the most to the indices, accounting for 1.32% of clinical visits (p-value = 0.02) and 3.55% of hospitalizations (p-value = 7.10×10^−9^). This may reflect the high prevalence of cardiometabolic diseases in the hospital-based TPMI cohort. In short, quantification of the impact of PRS for various diseases and traits on human health opens up opportunities for developing precision health management strategies.

## Discussion

This study represents a lagre-scale GWAS in the population of Han Chinese ancestry, utilizing data of around 500,000 individuals recruited from 16 medical centers across Taiwan. We investigated the genetic architecture of 695 dichotomized phecodes and 24 quantitative traits, identifying 2,656 independent variant-trait associations and showed that population-specific genetic risk-prediction PRS models for a wide range of diseases performed well in the population. Indeed, for the traits with sufficient sample size in the cohort, the PRS performance rival those developed for EUR using UKB data. These findings show that population-specific PRS models can be developed successfully for populations of non-European ancestry and our project serves as a model for large-scale genetic studies in other populations.

Recent large-scale projects that emphasize ancestral diversity in human genetic studies have discovered new findings with the inclusion of subjects of non-European ancestry. MVP conducted multi-ancestry GWAS on 635,000 participants, identifying over 2,000 signals unique to populations with non-European ancestry^16^. With the TPMI dataset, we performed larger GWAS in subjects of Han Chinese ancestry for several traits than published studies. For instance, the previous largest meta-analysis for type 2 diabetes included 20,573 cases who were of Han Chinese ancestry^37^. In contrast, our GWAS included 59,289 cases of type 2 diabetes, almost tripling the number of cases ever tested, and identified five unreported T2D SNPs from known regions, demonstrating the power of TPMI sample size. Identification of new and population-specific risk variants may lead to further understanding of their molecular mechanism and underline the need for population-specific weightings in PRS models. Moreover, population-specific findings also explain better performance of population-specific PRS model in the population in question. In short, our population-specific genomic profiles for comprehensive phenotypes provide a solid foundation for PRS development.

Our understanding of how the genetic factors influencing hepatitis B, an endemic infectious disease in Taiwan with an estimated hepatitis B virus carrier rate of 9.78% among the unvaccinated cohort (born before 1984)^25^, also benefited from the large dataset. With 23,618 cases, a significant increase from prior studies of only a few thousand cases^38–40^, we identified 26 fine-mapped signals, including 19 novel loci, and revealed a significant negative correlation between hepatitis B and other autoimmune diseases, such as Sicca syndrome, psoriasis, and systemic lupus erythematosus. Our well-performed and validated PRS model for hepatitis B demonstrated that host genome may determine the severity and symptoms of this infectious disease. This is similar to that previously reported in COVID-19 and pneumonia, where genetic factors have been shown to influence disease outcomes^41–44^. Our unexpected success of GWAS and PRS for hepatitis B not only demonstrate the power of the large sample size of TPMI, but also reveal the necessity of population-specific genetic study for population-enriched diseases. The benefits extend beyond differences in ancestry to include environmental factors such as pathogen exposure, food intake, and lifestyle influences. Exploring how the human genome interacts with these diverse external and environmental factors can significantly enhance our understanding of how genetic variants contribute to disease susceptibility or severity.

In addition, the comprehensive phenotypic data allow us to investigate the genetic correlation among multiple traits, which have substantial implications for clinical applications, and leverage them to improve the performance of PRS models. By identifying shared genetic risks across diseases, at-risk individuals can be alerted to pursue early detection of comorbidities and targeted prevention strategies. For example, the clustering of cardiometabolic traits, such as type 2 diabetes, hypertension, and BMI, highlights their interconnected genetic basis and suggests that individuals with a high genetic risk for one condition may benefit from early screening and intervention for related conditions^45^. Additionally, the shared genetic architecture allows the development of multi-trait PRS models that integrate genetic risks across correlated traits, improving prediction accuracy and enabling precision medicine approaches that address multiple health outcomes simultaneously^46^. Including the correlated traits in PRS model development improved performance, resulting in an average 1.55-fold increase in the explained percentage of phenotypic variation (p-value = 1.07×10^−13^). Although previous studies have proven the utility of multi-traits on target diseases^36,47,48^, we have extended the use this approach on a phenome-wide level and demonstrated the improvement across different types of traits. As a result, we produced well-performed PRS for various categories of diseases, including cardiometabolic diseases, autoimmune disorders, and infectious diseases.

We evaluated our PRS models across several large cohorts, including the TWB, UK Biobank, and All of Us. The TPMI-derived PRS models consistently outperformed those developed from European populations when applied to diseases in Han Chinese descents or EAS from the three large cohorts. When comparing with EUR-derived PRS models, we also observed better performance across several traits in EAS, particularly for cardiometabolic and autoimmune diseases. Similarly, the TPMI-included cross-population model slightly improves performance in populations of other ancestries. These results highlight the need for population-specific models and emphasize the importance of genetic data from diverse populations to advance cross-population models. By integrating these well-developed PRS models, we estimate that genetics account for 10.3% of variation of hospitalization duration in TPMI. Although the estimates of genetic contributions to health measure may be influenced by disease prevalences and ascertainment biases, our result suggest integrating genetic risk-based health management strategies with traditional risk factors, such as age, sex, smoking, and BMI, may enhance prediction models and refine personalized risk stratification.

As with other large-scale epidemiological studies, ascertainment bias is also observed in our study. TPMI’s case proportion shows a significant but moderate correlation with the prevalence from NHIRD, implying potential ascertainment bias of TPMI’s hospital-based design. Compared to the general population in Taiwan (NHIRD), TPMI participants are overrepresented in the middle-aged group (year of birth 1940-1970, 54.3% vs. 38.3%), include slightly more females (55.1% vs. 50.6%), and have a higher proportion of participants from northern Taiwan (59.5% vs. 47.3%). These demographic differences, along with the volunteer-based recruitment process, likely contribute to the lower case proportions observed in TPMI (Figure 1a). Importantly, a significant portion of disease records in the NHIRD originate from local clinics and primary care settings, which are not covered in TPMI. Notably, the EMR of the participants are incomplete, as some participants receive care from multiple health providers, but the TPMI only has access to EMRs from their enrollment hospitals. We acknowledge that ascertainment biases may influence disease prevalence and heritability estimates. Thus, we accounted for case-control ascertainment by applying liability-scale transformations using population prevalence data from NHIRD and utilized independent validation for PRS (TWB, UKB, All of Us) to mitigate the impact of ascertainment biases, while acknowledging that residual biases may persist. Methods like inverse probability weighting could mitigate such biases^49^, but these require detailed external reference data that are currently unavailable. Additionally, we observed a relatively low estimated heritability for body height and BMI in TPMI, compared to values reported in the literature.^50,51^ These estimates may be affected by factors such as inconsistencies in assessment across EMR, variations in statistical approaches, and reduced bias of assortative mating in TPMI population.^52–54^ These limitations also emphasise the need for future adjustments to enhance generalizability.

In addition to the ascertainment bias, our study has other commonly found limitations. First, the TPMI cohort size is not sufficiently large to study some of the severe subtypes of many diseases, such as diabetes insipidus and neurofibromatosis. Second, we attempted to use eQTLs to elucidate the molecular mechanism of diseases, but the underrepresentation of EAS in current eQTL datasets, such as GTEx, poses challenges^27^. Gene expression regulation varies across ancestries^55,56^, and differences in LD structures further complicate colocalization analyses. Comparing to GTEx whole blood eQTL, the multi-ancestry lymphoblastoid eQTL and Japanese whole blood eQTL revealed 309 additional gene-trait pairs. Therefore, ancestral diversity is an urgent need not only in genomic data but also in transcriptomic, proteomic, metabolomic, and epigenomic datasets. Third, the current project retrieved EMRs from an average of 5 years prior to enrollment, so some important data such as age of disease onset for the older participants are not available. Incomplete EMR leads to less precise case definition of some participants. Fourth, some of the younger participants have high-risk genetic profiles but are disease free for those diseases. The duration of the project is too short to determine whether they will eventually develop those diseases.

Effort is underway to gain access to the complete EMRs of the TPMI participants and to recruit additional participants with severe subtypes of common diseases. The high-risk participants who are symptom-free are being followed to monitor disease development. Future studies are being planned to study the high-risk individuals who escape disease development to identify genetic and non-genetic factors that mitigate their disease risk. Furthermore, the meta-analysis integrating TPMI with other large-scale EAS biobanks, such as TWB^11,12^, Korean Genome and Epidemiology Study^9^, China Kadoorie Biobank^10^, and Biobank Japan^8^, may further enhance our understanding of the genetic etiology in EAS and improve prediction models.

This study demonstrates that population-specific risk-prediction models, such as those developed for EAS in this work, can achieve strong predictive performance for traits with high relevance in that population. The PRSs we developed for EAS performed well for multiple traits, including diseases with significant public health implications, such as type 2 diabetes and systemic lupus erythematosus. However, for certain traits, such as female breast cancer and glaucoma, PRS derived from both UK Biobank and TPMI performed comparably, suggesting that the genetic architecture of some traits allows for generalizable models. These findings emphasize the importance of developing and validating PRS models in diverse global populations to maximize their utility and equity in genetic risk prediction. While our results emphasize the utility of developing population-specific PRS, further research is needed to directly compare their performance with multi-population models and assess their generalizability and to assess their impact on disease prevention and management. In particular, longitudinal studies and real-world implementations will be critical to determine the extent to which PRS-guided interventions can delay disease onset or improve health outcomes. Furthermore, it is hopeful that if all can obtain their genetic profiles and determine their risk for major diseases, many diseases can be prevented or their onset can be delayed significantly, thereby fulfilling the promise of modern genetics.

In conclusion, we used a large-scale Han Chinese dataset produced by the TPMI to conduct pheno-wide genetic analyses and leverage these genetic findings to train risk prediction models for multiple diseases and traits. The developed models are validated in EAS of different biobanks and demonstrate a consistent performance that bodes well for their use in the general Han Chinese/EAS population. Our approach can serve as a template for developing PRS models in populations currently without such resources, anticipating the time when all populations around the world can benefit from risk-based health management as part of the precision health movement.

## Methods

### Study Population and Phenotyping

We utilized the Taiwan Precision Medicine Initiative (TPMI) dataset, which links extensive electronic medical records (EMR) with genotypic data for 486,956 individuals. Dichotomized disease status was defined by phecodes, which were based on information extracted from the EMR using International Classification of Diseases (ICD) codes^18,19^. To ensure robustness, cases were defined by having the diagnosis of the relevant condition on two or more clinical visits. We also extracted quantitative traits from the EMR, including anthropometric, vital sign and laboratory measurements, and we excluded the extreme outliers and removed or adjusted the treated and/or medicated measures based on previous research, and the median value was kept if the participant had multiple qualified measures^59^. (Supplementary methods) In this study, we focused on 695 dichotomized phenotypes (phecodes) that had at least 2,000 cases and 24 quantitative traits that were measured in at least 100,000 individuals. These phecodes spanned 17 disease categories, including but not limited to infectious diseases, neoplasms, endocrine/metabolic disorders, and circulatory system diseases. The 24 quantitative traits were categorized into anthropometric, circulatory, hematological, kidney-related, liver-related, and metabolic measurements.

### Genotyping and Quality Control

Genotyping was performed using two customized high-density Axiom SNP arrays produced by Thermo Fisher (Waltham, MA, USA), TPMv1 and TPMv2. The genotyping experiments were conducted in six genotyping centers in Taiwan^60^. The raw genotypic data underwent quality control measures, and the genetic variants were excluded when they had call rate <0.98 minor allele frequency (MAF) <0.01, or Hardy-Weinberg equilibrium test p-value <1×10^−6^. We also excluded individuals with overall call rate <0.95, failed heterozygosity check, or inconsistent documented versus genetically determined sex. For this study, we only included the genetic variants found on both genotyping arrays and excluded variants with a significant batch effect in GWAS. The proportion of genetic ancestry was determined by ADMIXTURE^61^, and the projected principal component scores with 1000 Genome as reference panel were applied to determine individuals’ ancestry^62^. As a result, 401,710 genetic variants and 463,447 Han Chinese participants passed all quality control measures and were used in the subsequent studies. Details are found in supplementary methods and GitHub (https://github.com/TPMI-Taiwan/tpmi-qc).

### Phasing and Imputation

Phasing was conducted on QC-passed genotype data with SHAPEIT5^63^. Genome imputation was carried out with IMPUTE5 using a reference panel of 1,498 whole genome sequenced Taiwan Biobank subjects^12,64^. We also conducted post-imputation quality control with exclusion criteria INFO score ≤0.7 and MAF ≤ 0.01. In addition, we also performed a chip-GWAS for minimizing the bias from different chips, resulting in a dataset of 8,046,864 well-imputed common genetic variants.

### Population Structure and Relatedness Estimation

We performed a principal component analysis (PCA) based on genotyped variants to capture the effect of population structure. To diminish the effect of close relatives, the main PCA was conducted in a genetically unrelated subset, and other subjects were projected with the calculated PC weightings. And then, these PCA scores were leveraged to accurately quantify the proportion of identity-by-decent (IBD) and degree of relatedness. The maximum unrelated set was determined based on these estimated degrees of relatedness. PC-AiR and PC-Relate were used for PCA and relatedness estimation and PRIMUS was used for identifying the maximum unrelated set with the third degree as threshold^65–67^.

### Genome-Wide Association Study (GWAS)

The entire dataset was divided into three subgroups: the GWAS set (N = 363,447), the training set (N = 80,000), and the testing set (N = 20,000). To maximize the statistical power, we used a mixed-effect regression model to examine the association between genotype and outcome of interest, logistic regression for dichotomized phecode, and linear regression for quantitative traits. The quantile-normalization was applied to quantitative traits to ensure the normal distribution. The mixed-effect model accounted for relatedness among individuals by including a random effect for pairwise kinship. The model also adjusted for key covariates, including age, sex, age^2^, interactions between age/age^2^ and sex, genotyping chip, enrollment hospital, and 10 genetic principal components to control for population stratification. SAIGE was applied for the mixed effect model GWAS^68^. Within the GWAS set, we selected an unrelated subset (N = 248,754) to perform GWAS using a generalized linear model with PLINK2, and we conducted 1:10 age, sex-matching for the traits with imbalanced case/control ratio (<1/20). These GWAS statistics were then used for heritability and genetic correlation estimation^69^.

### Replication Evaluation

To systematically evaluate the performance of our GWAS, we leveraged a pre-summarized phenotype-genotype reference map^70^, which collected 5,879 genetic associations for 149 unique phecodes from 523 published GWAS and 1,215 associations from EAS. We calculate the overall and power-adjusted replication rates and actual over expected ratio for each available phecode and categories respectively. For measuring the quality of biobank data through replication an R package PGRM was used^70^.

### Fine-mapping

We performed fine-mapping to identify the independent GWAS signals in all genomic regions containing any variant with a p-value<5×10^−8^ and ±1.5 Mb of the regional lead variant^14^, except the major histocompatibility complex region (MHC region, chr6: 25,391,792-33,424,245) due to its complex LD structure. We used the reported 95% credible set to determine the independent signals, and up to ten signals were allowed for each region. The genome-wide significant threshold was applied for defining a credible set as an independent hit, and an additional requirement of log Bayes factor (BF) > 2 was applied for the second hit. For the failed fine-mapping regions and MHC region, we used the lead SNP as the hit of each significant region. SuSiE was conducted for this summary statistics-based fine-mapping with LD derived from our imputation reference panel^71^, which reflects our study population’s genetic architecture. While using LD from the GWAS sample might improve accuracy, we used the imputation panel due to the computation efficiency.

### Novel Association Identification

We comprehensively compared our GWAS results with reported significant signals on the NHGRI-EBI GWAS Catalog^21^, download at 2024/03/11. The mapping of phecodes and quantitative traits to GWAS catalog phenotypes is summarized in Table S16. We classified a variant as novel if the fine mapped independent signal was not located within 1 Mb of any reported genome-wide significant association (p-value<5×10^−8^) for the corresponding phenotype. Additionally, a variant was considered a new hit if the highest linkage disequilibrium (LD) r^2^ was less than 0.1 with any reported significant association within 1Mb. Associations derived from uncertain and umbrella phecodes were excluded, and for duplicated genetic variants or regions, we only reported the association with the smaller p-value or from the phecode with the more specific definition. Finally, we used ANNOVAR to annotate the novel variants with data from the RefSeq Gene database (2020-08-17 updated)^72,73^. For the novel variants, we explored their allele frequencies in EUR (non-Finnish), AFR, SAS, AMR from gnomAD^74^. We also compared the the effect size between TPMI and UKB with t-test for investigating the ancestry-specific effect.

### Heritability, Genetic Correlation, and Clustering

To quantify the genomic contribution of the specific traits, we applied linkage disequilibrium score regression to estimate the SNP-based heritability with LDSC^26^. The GWAS summary statistics and the pre-calculated LD score from EAS superpopulation of 1000 Genome were used^62^. For the dichotomized traits, we performed a liability-scaled transformation on the observed heritability using the 5-years population prevalence from the National Health Insurance dataset (NHIRD) of the Health and Welfare Data Science Center^20,75^. For the traits with a higher prevalence in our dataset (TPMI) than the population (NHIRD), we applied the equation derived by Lee et al^76^. For other traits, we used the adapted equation from Ojavee et al^75^. Additionally, we conducted LDSC to obtain pairwise genetic correlations to assess the similarity of genetic mechanisms between traits^77^. Based on the genetic correlation matrix, we used a hierarchical cluster analysis to identify groups of traits that share genetic mechanisms. We employed the weighted pair group method with arithmetic mean (WPGMA) for clustering, and the resulting cluster tree was used for group identification. Moreover, we estimate the genetic correlation across populations, TPMI and UK biobank, to demonstrate varied genetic architecture in different ancestry populations. For the UK biobank GWAS, we applied a generalized linear model from PLINK2 with the predefined phecode, https://github.com/umich-cphds/createUKBphenome, and corresponding baseline quantitative measures among the identified unrelated set (n = 378,544). Popcorn was performed for the cross-population genetic correlation, and two correlation coefficients were calculated, the transethnic genetic-effect correlation (ρ_ge_) and transethnic genetic-impact correlation (ρ_gi_)^30^.

### Gene-Level Heritability and Colocalization

We used both gene-level heritability estimation and colocalization analysis to map our GWAS findings to functional units, specifically genes. We conducted h2gene analysis to partition SNP-based heritability to the gene level^57^. We estimated heritability for genes that overlapped with fine-mapped regions, where gene regions were defined as the gene body ±10 kb for gene-level heritability. Additionally, to illustrate the molecular functions of genes of interest, we used colocalization analysis to examine whether there are shared common genetic causal variants between tissue-specific gene expression and traits of interest. We utilized expression quantitative traits locus (eQTL) resources from 49 tissues in GTEx v8^27^, lymphoblastoid in MAGE^28^, and whole blood in JCTF^29^, testing any gene with genome-wide significant signals in the cis-regulation region (±1 Mb). The posterior probabilities were used to evaluate colocalization between gene expression and the trait of interest. The R package, coloc, was used with SuSiE relaxing the single causal variant assumption^58,78,79^.

### Single and Multi-Trait Polygenic Risk Score (PRS)

The preserved dataset of 100,000 unrelated TPMI subjects was split into two subsets, training (n = 80,000) and validation (n = 20,000) for PRS model building. Five popular PRS tools were used, LDpred2^31^, Lassosum2^32^, PRS-CS^33^, SBayesR^34^, and MegaPRS^35^, and the training subset was applied for parameter selection and model optimization if needed. LDpred2, PRS-CS, and SBayesR assumed the effect of genetic variants following a mixture distribution with different pre-defined parameters and applied a Bayesian framework for distribution estimation. Lassosum2 utilized a penalized regression (LASSO) for weight estimating, and MegaPRS leveraged minor allele frequency and linkage disequilibrium for model building. We then used the validation subset to evaluate the performance of PRS models. Individual score was calculated with PLINK2^69^. The explained variance (r^2^) was used to evaluate the performance of PRS for quantitative traits^76,80^, and two indices, area under the receiver operating characteristic curve (AUC) and liability-scaled r^2,^ were used for PRS of dichotomized phenotypes. We followed the approach by Ni et al.^80^ and report both raw r^2^ and r^2^ adjusted for covariates (sex, age, and PCs). Additionally, we include partial r^2^ estimates, calculated using the R package rsq. To account for population stratification in cross-cohort predictions, we also report r^2^ with PCs as covariates in the supplemental tables. For AUC comparisons, we include a baseline model incorporating standard covariates (sex, age, and PCs) to better assess the added predictive power of PRS. The likelihood ratio test was used to obtain the significance for r^2^ with R package, lmtest, and standard error for AUC was calculated with R package, auctestr. To further leverage the gene’s pleiotropy and shared genetic mechanism among traits, we conducted a multi-trait PRS model building for the traits in the same genetic cluster based on pairwise genetic correlation identified in the previous step. We pooled all PRS models from five tools for those identified traits and applied an elastic net regression to combine their weighting and find the most optimized model for the target trait. PRSmix+ was performed for the multiple traits PRS model building^36^. The cross-population PRS models was based on both TPMI and UKB European GWAS, and PRS-CSx was applied^81^.

### External Validation and Comparison

We conducted an external validation of our developed PRS using data from the Taiwan Biobank, EAS from UK Biobank and All of Us. TWB is a community-based biobank, and it has recruited over 200,000 participants in Taiwan. Herein, we used 88,628 unrelated subjects (> third degree and removed 5,242 overlapped individuals), who were genotyped with the Axiom customized chip TWB2 (equivalent to TPMv1), and their genotyping QC, phasing, and imputation followed the same protocol as described above. The self-reported disease condition was queried from their baseline questionnaire, except for cancer. Since the study design of TWB excluded cancer patients at recruitment, we used both baseline and follow-up self-reporting data to define cancer cases and controls. UKB has enrolled ~500,000 participants since 2006 and linked their genetic data with enriched phenotypic data. For UKB validation, we used their inpatient record for case definition. Their ancestral population was determined by self-reported ethnic background, such as self-reported Chinese as EAS (n = 1,572), White, British, Irish, and any other white background as EUR (n = 472,869), Black or Black British, Caribbean, African, and any other Black background as AFR (n = 8,074), and Asian or Asian British, Indian, Pakistani, Bangladeshi, and any other Asian background as SAS (n = 9,893). All of Us intends to enroll over 1 million participants in the United States and has released whole genome genotyping data for ~312,000 participants as of the first quarter of 2024. We applied ADMIXTURE with 1000 Genomes as reference panel for assigning the genetically inferred ancestral populations, including EAS (n = 6,895), EUR (n = 152,754), AFR (n = 60,964), AMR (n = 32,394), and SAS (n = 2,334). The genetically confirmed EAS as well as other superpopulations and their linked EMR were used for validating our PRS models. Moreover, we compared the TPMI-derived PRS model with UKB-derived models to investigate the performance of population-specific PRS. The UKB-derived models were based on published UKB European GWAS (https://pheweb.org/UKB-TOPMed/), and LDpred2-auto was applied for model building.

### Overall Health Measures Evaluation

We evaluated the genetic impact on overall health meaures. We used the number of clinical visits and the aggregate duration of hospitalization as overall health indices. Due to collinearity among PRS for different traits, we utilized a partial least square-generalized linear model (PLS-GLM) to extract components from the PRS of qualified traits with R package, plsRglm^82^. The number of extracted components was determined by the Akaike Information Criterion (AIC). We then estimated the covariate-adjusted proportion of genetic contribution (r^2^) by comparing the full model with the null model, which included only covariates such as sex, age, and hospital. Likelihood ratio test was used to obtain the significances of regression models. For each index, we employed three models to compare the top and bottom 5%, 10%, and 20%. We selected covariate-matched controls from subjects without hospitalization records as the bottom group for hospitalization models.

## Code availability

Code for genotyping quality control process and analysis is available at our Github (https://github.com/TPMI-Taiwan/).

## Data availability

All polygenic risk score (PRS) models and GWAS results (summary statistics from SAIGE and PLINK) are available from the TPMI website. The UKB phecode GWAS was obtatined from UKBiobank TOPMed-imputed PheWeb (https://pheweb.org/UKB-TOPMed/). The eQTL resources are download from websites of GTEx (https://www.gtexportal.org/), MAGE (https://doi.org/10.5281/zenodo.10535719), and JCTF (https://humandbs.dbcls.jp/en/hum0343-v4). The detailed describtion of data availability and application process of TWB, NHIRD, UKB and All of Us can be found on their websites (TWB: https://www.biobank.org.tw/english.php; NHIRD: https://www.apre.mohw.gov.tw/; UKB: https://www.ukbiobank.ac.uk/; All of Us: https://www.researchallofus.org/).

## Ethics

This study was approved by the Institutional Review Boards of Taipei Veterans General Hospital (2020-08-014A), National Taiwan University Hospital (201912110RINC), Tri-Service General Hospital (2-108-05-038), Chang Gung Memorial Hospital (201901731A3), Taipei Medical University Healthcare System (N202001037), Chung Shan Medical University Hospital (CS19035), Taichung Veterans General Hospital (SF19153A), Changhua Christian Hospital (190713), Kaohsiung Medical University Chung-Ho Memorial Hospital (KMUHIRB-SV(II)-20190059), Hualien Tzu Chi Hospital (IRB108-123-A),Far Eastern Memorial Hospital (110073-F), Ditmanson Medical Foundation Chia-Yi Christian Hospital (IRB2021128), Taipei City Hospital (TCHIRB-10912016), Koo Foundation Sun Yat-Sen Cancer Center (20190823A), Cathay General Hospital (CGH-P110041), Fu Jen Catholic University Hospital (FJUH109001) and Academia Sinica (AS-IRB01-18079), Taiwan. Written informed consent was obtained from the subjects in accordance with institutional requirements and the Declaration of Helsinki principles. All collected information was de-identified before statistical data analysis. The analysis with Taiwan Biobank was apporaved by Institutional Review Boards of Academia Sinica (AS-IRB-BM-19014), and the NHIRD analysis with Health and Welfare Data Science Center (HWDC) was approved by Institutional Review Boards of Academia Sinica (AS-IRB-BM-23056). This research has been conducted using the UK Biobank Resource under UK Biobank Main Application 15326. Work with All of Us data was performed using the All of Us Researcher Workbench under the workspace “Duplicate of Prediction of Polygenic Traits’”.

## Acknowledgements

We thank all the participants and researchers of the Taiwan Precision Medicine Initiative and the Taiwan Biobank. We gratefully acknowledge the use of data from the National Health Insurance Research Database, provided by the Ministry of Health and Welfare of Taiwan. This study was funded in part by the Academia Sinica (40-05-GMM, AS-GC-110-MD02, and 236e-1100202 to P.-Y.K. and J.-Y.W.) and the National Development Fund, Executive Yuan (NSTC 111-3114-Y-001-001 to P.-Y.K.). This work used ASGC (Academia Sinica Grid-computing Center) Distributed Cloud resources, which is supported by Academia Sinica. Analysis using UK Biobank data used computational resources hosted by the Michigan State University High-Performance Computing Center. Data from the UK Biobank includes data provided by patients and collected by the National Health Service (NHS) England as part of their care and support. UK Biobank data also includes data assets made available by National Safe Haven as part of the Data and Connectivity National Core Study, led by Health Data Research UK in partnership with the Office for National Statistics and funded by UK Research and Innovation (research which commenced between 1st October 2020--31st March 2021 grant ref MC_PC_20029; 1st April 2021--30th September 2022 grant ref MC_PC_20058). Also, we are grateful and acknowledge the contributions of the All of Us participants who make this project possible and the work of the National Institutes of Health’s All of Us Research Program for making this data available.

## Author contribuctions

Supervision: P.-Y.K., W.H.S., S.-F.Y., J.-M.L., J.-Y.W., J.-F.C., J.-Y.W.,, C.S.F., M.-C.H., Y.-C.F., and C.-H.C.; Formal analysis: H.-H.C., C.-Y.C., J.-P.S., W.-J.L., F.-J.H., E.H.W., E.-C.Y., S.-H.W., and S.-P.C.; Resources: P.-Y.K., C.-L.C., J.-K.J., I.-H.L., K.-H.L., W.-S.C., H.-C.T., S.-Y.L., F.-P.C., H.-L.H., Y.-C.Y., W.-C.T., M.-H.L., H.-T.C., L.-M.T., W.-Y.L., P.C.C., Y.-C.H., Y.-M.C., T.-H.H., C.-H.L., Y.-J.C., I.-C.C., C.-L.M., S.-J.C., Y.-L.C., Y.-J.L., C.-H.L., W.-J.L., H.T., T.-T.Y., H.-C.Y., M.-Y.C., Y.-C.L., Y.-T.K., B.-Z.K., J.-E.L., C.-L.C., J.-C.L., P.C., C.-H.L., C.-H.C., I.-C.W., L.-C.L., J.-W.W., S.-l.S., S.-W.H., C.-H.H., W.-M.L., C.-J.Y., Y.-T.C., D.-R.H., Y.-H.H, C.-H.Y., Y.-S.H., Y.-F.C., H.-M.W., P.-H.T., K.-G.H., C.-Y.C., Y.-L.H., M.-S.W., J.-H.K., Y.-B.L., J.J.J., Y.-H.L., J.-Y.L., H.-J.L., A.-C.F., J.-S.L., N.-F.C., Y.-W.Y., E.M., C.-Y.H., C.-C.L., T.-F.W., K.-Y.S., J.-K.W., M.-H.C., H.-F.C., G.-C.M., T.-Y.C., F.-T.C., H.-J.C., K.-J.K., C.-F.H., C.-Y.T., P.-Y.C., K.T. and Y.-J.S.; Validation (TWB): H.-H.C., C.-Y.C., J.-P.S., W.-J.L., F.-J.H., E.H.W., E.-C.Y., and J.-Y.W.;Validation (UKB and All of Us): T.R., E.W., S.H.; Data Curation:M.-F.T., T.-H.W., Y.-C.L.; Writing - Original Draft: H.-H.C., C.S.F., L.-H.L., C.-Y.C., J.-P.S., W.-J.L., E.H.W., E.-C.Y.; Writing - Review & Editing: P.-Y.K., E.-C.Y., M.-F.T., C.-H.C., H.-C.Y., Y.-T.H., C.-H.C., C.-Y.W., H.-H.C., C.S.F., and L.-H.L.; Investigation: L.-H.L., R.-H.W., Y.-C.C., and C.-P.C.; Project administration: Y.-M.L. and C.-S.Y. All authors reviewed and approved the final manuscript.

## Extended Data

**Extended Data Fig. 1**. Scatter plots of the case proportion for dichotomized phenotypes by disease categories. The significantly positive correlations are observed cross disease categories between the case proportion in TPMI (x-axis) and the 5-ear prevalence in NHIRD (y-axis), except congenital anomalies.

**Extended Data Fig. 2**. Comparison of TPMI GWAS-identified loci to the previously published GWAS. The replication rates of TPMI GWAS-identified loci when compared previously reported loci from the GWAS catalog are presented in this bar chart. Red ars indicate the comparison of TPMI findings to that of all ancestries and blue bars present the comparison to East Asian ancestries. The categories of diseases are hown under the bars.

**Extended Data Fig. 3**. The Manhattan plot of GWAS for viral hepatitis B in TPMI. The names of nearest mapped gene were labeled for the independent GWAS significant ci.

**Extended Data Fig. 4**. Genetic correlation heatmap for all heritable traits. Heatmap showing genetic correlations among heritable traits. Genetic correlations were estimated using LDSC, with colors representing the correlation coefficients between traits. The weighted pair group method with arithmetic mean (WPGMA) was used for clustering with the correlation coefficient as distance between traits.

**Extended Data Fig. 5**. The scatter plot of performance for PRS developed by different tools. The color represents the phecode category, and the shape indicates the PRS development tool: Lassosum2 (circle), LDpred2 (triangle), MegaPRS (square), PRS-S (cross), and SbayesR (square cross). Each phecode is positioned at a unique x-coordinate, with the tool that has the highest AUC highlighted.

**Extended Data Fig. 6**. The bar chart and dot plot for PRS performance. Bar and dot plot showing PRS explained variance (r^2^) and SNP-heritability for dichotomous traits. Gray bars indicate SNP-heritability (estimated from TPMI GWAS unrelated set [n =248,754] with LDSC), and the colored bar chart presents the r^2^ values, indicating the proportion of variance explained by the PRS among TPMI validation set (n =20,000), and dots and error bars show Area Under the receiver operating characteristic Curve (AUC) with 95% confidence interval. An asterisk (*) indicates estimates considering the MHC region.

**Extended Data Fig. 7**. External validation of PRS models in Taiwan Biobank and other cohorts. PRS performance is presented as Area Under the receiver operating characteristic Curve (AUC) with 95% confidence inverval in TPMI (orange, TPMI validation set, n = 20,000), Taiwan Biobank (green, n = 88,628), East Asians in UK Biobank (blue, n = 1,572), and East Asians in All of Us (purple, n = 6,895). Circles represent TPMI-derived PRS, and triangles indicate UKB (European)-derived PRS models. Only the estimates with case size > 30 were showed on the figure.

**Extended Data Fig. 8**. External validation of TPMI-derived PRS model and TPMI-UKB cross-population PRS models across populations. The plots show the Area Under the receiver operating characteristic Curve (AUC) with 95% confidence inverval for PRS validation in East Asian (red, n = 88,628 in TWB; 1,572 in UKB; 6,895 in All of Us), European (olive green, n = 472,869 in UKB and 152,754 in All of Us), African (green, = n = 8,074 in UKB and 60,964 in All of Us), South Asian (blue, n = 9,893 in UKB and 2,334 in All of Us), and Admixed American (purple, n = 32,394 in All of Us) populations om TPMI (circle), UKB (triangle), and All of Us (square) cohorts.

**Extended Data Table 1**. Proportion of overall health indices explained by genetic risk ^1^Model adjusting for sex, age and enrollment hospital

Likelihood ratio test was applied to obtain the explained variation (r2) and significance y comparing the full model and null model.

## Supplementary Information

### Supplemental tables

Table S1-S16

### Supplemental text

Detailed methods of genotyping and phenotyping quality control, and SNP-based heritability for quantitative traits

## References

1 Lennon, N. J. et al. Selection, optimization and validation of ten chronic disease polygenic risk scores for clinical implementation in diverse US populations. Nat Med 30, 480–487 (2024). 10.1038/s41591-024-02796-z

2 Thompson, D. J. et al. A systematic evaluation of the performance and properties of the UK Biobank Polygenic Risk Score (PRS) Release. PLoS One 19, e0307270 (2024). 10.1371/journal.pone.0307270

3 Mills, M. C. & Rahal, C. The GWAS Diversity Monitor tracks diversity by disease in real time. Nat Genet 52, 242–243 (2020). 10.1038/s41588-020-0580-y

4 Duncan, L. et al. Analysis of polygenic risk score usage and performance in diverse human populations. Nat Commun 10, 3328 (2019). 10.1038/s41467-019-11112-0

5 Martin, A. R. et al. Human Demographic History Impacts Genetic Risk Prediction across Diverse Populations. Am J Hum Genet 100, 635–649 (2017). 10.1016/j.ajhg.2017.03.004

6 Bentley, A. R., Callier, S. & Rotimi, C. N. Diversity and inclusion in genomic research: why the uneven progress? J Community Genet 8, 255–266 (2017). 10.1007/s12687-017-0316-6

7 Smith, J. L. et al. Multi-Ancestry Polygenic Risk Score for Coronary Heart Disease Based on an Ancestrally Diverse Genome-Wide Association Study and Population-Specific Optimization. Circulation: Genomic and Precision Medicine 17, e004272 (2024). 10.1161/CIRCGEN.123.004272

8 Ishigaki, K. et al. Large-scale genome-wide association study in a Japanese population identifies novel susceptibility loci across different diseases. Nat Genet 52, 669–679 (2020). 10.1038/s41588-020-0640-3

9 Nam, K., Kim, J. & Lee, S. Genome-wide study on 72,298 individuals in Korean biobank data for 76 traits. Cell Genom 2, 100189 (2022). 10.1016/j.xgen.2022.100189

10 Walters, R. G. et al. Genotyping and population characteristics of the China Kadoorie Biobank. Cell Genom 3, 100361 (2023). 10.1016/j.xgen.2023.100361

11 Feng, Y. A. et al. Taiwan Biobank: A rich biomedical research database of the Taiwanese population. Cell Genom 2, 100197 (2022). 10.1016/j.xgen.2022.100197

12 Wei, C. Y. et al. Genetic profiles of 103,106 individuals in the Taiwan Biobank provide insights into the health and history of Han Chinese. NPJ Genom Med 6, 10 (2021). 10.1038/s41525-021-00178-9

13 Sudlow, C. et al. UK biobank: an open access resource for identifying the causes of a wide range of complex diseases of middle and old age. PLoS Med 12, e1001779 (2015). 10.1371/journal.pmed.1001779

14 Kurki, M. I. et al. FinnGen provides genetic insights from a well-phenotyped isolated population. Nature 613, 508–518 (2023). 10.1038/s41586-022-05473-8

15 All of Us Research Program Genomics Investigators. Genomic data in the All of Us Research Program. Nature 627, 340–346 (2024). 10.1038/s41586-023-06957-x

16 Verma, A. et al. Diversity and scale: Genetic architecture of 2068 traits in the VA Million Veteran Program. Science 385, eadj1182 (2024). 10.1126/science.adj1182

17 Yang, H.-C. et al. The Taiwan Precision Medicine Initiative: A Cohort for Large-Scale Studies. bioRxiv, 2024.2010.2014.616932 (2024). 10.1101/2024.10.14.616932

18 Bastarache, L. Using Phecodes for Research with the Electronic Health Record: From PheWAS to PheRS. Annu Rev Biomed Data Sci 4, 1–19 (2021). 10.1146/annurev-biodatasci-122320-112352

19 Denny, J. C. et al. Systematic comparison of phenome-wide association study of electronic medical record data and genome-wide association study data. Nat Biotechnol 31, 1102–1110 (2013). 10.1038/nbt.2749

20 Lin, L. Y., Warren-Gash, C., Smeeth, L. & Chen, P. C. Data resource profile: the National Health Insurance Research Database (NHIRD). Epidemiol Health 40, e2018062 (2018). 10.4178/epih.e2018062

21 Sollis, E. et al. The NHGRI-EBI GWAS Catalog: knowledgebase and deposition resource. Nucleic Acids Res 51, D977–D985 (2023). 10.1093/nar/gkac1010

22 Zhou, Y. et al. Performance of multigene testing in cytologically indeterminate thyroid nodules and molecular risk stratification. PeerJ 11, e16054 (2023). 10.7717/peerj.16054

23 Tyagi, A., Goyal, A., Chaware, P. & Rathinam, B. A. D. Mutations of PHOX2B Gene in Patients of Obesity Hypoventilation Syndrome in Central India. J Lab Physicians 14, 164–168 (2022). 10.1055/s-0041-1735582

24 He, D. et al. A longitudinal genome-wide association study of bone mineral density mean and variability in the UK Biobank. Osteoporos Int 34, 1907–1916 (2023). 10.1007/s00198-023-06852-1

25 Chang, K. C. et al. Survey of hepatitis B virus infection status after 35 years of universal vaccination implementation in Taiwan. Liver Int 44, 2054–2062 (2024). 10.1111/liv.15959

26 Bulik-Sullivan, B. K. et al. LD Score regression distinguishes confounding from polygenicity in genome-wide association studies. Nat Genet 47, 291–295 (2015). 10.1038/ng.3211

27 GTEx Consortium. The GTEx Consortium atlas of genetic regulatory effects across human tissues. Science 369, 1318–1330 (2020). 10.1126/science.aaz1776

28 Taylor, D. J. et al. Sources of gene expression variation in a globally diverse human cohort. Nature 632, 122–130 (2024). 10.1038/s41586-024-07708-2

29 Wang, Q. S. et al. The whole blood transcriptional regulation landscape in 465 COVID-19 infected samples from Japan COVID-19 Task Force. Nat Commun 13, 4830 (2022). 10.1038/s41467-022-32276-2

30 Brown, B. C., Asian Genetic Epidemiology Network Type 2 Diabetes, C., Ye, C. J., Price, A. L. & Zaitlen, N. Transethnic Genetic-Correlation Estimates from Summary Statistics. Am J Hum Genet 99, 76–88 (2016). 10.1016/j.ajhg.2016.05.001

31 Prive, F., Arbel, J. & Vilhjalmsson, B. J. LDpred2: better, faster, stronger. Bioinformatics 36, 5424–5431 (2021). 10.1093/bioinformatics/btaa1029

32 Prive, F., Arbel, J., Aschard, H. & Vilhjalmsson, B. J. Identifying and correcting for misspecifications in GWAS summary statistics and polygenic scores. HGG Adv 3, 100136 (2022). 10.1016/j.xhgg.2022.100136

33 Ge, T., Chen, C. Y., Ni, Y., Feng, Y. A. & Smoller, J. W. Polygenic prediction via Bayesian regression and continuous shrinkage priors. Nat Commun 10, 1776 (2019). 10.1038/s41467-019-09718-5

34 Lloyd-Jones, L. R. et al. Improved polygenic prediction by Bayesian multiple regression on summary statistics. Nat Commun 10, 5086 (2019). 10.1038/s41467-019-12653-0

35 Zhang, Q., Prive, F., Vilhjalmsson, B. & Speed, D. Improved genetic prediction of complex traits from individual-level data or summary statistics. Nat Commun 12, 4192 (2021). 10.1038/s41467-021-24485-y

36 Truong, B. et al. Integrative polygenic risk score improves the prediction accuracy of complex traits and diseases. Cell Genom 4, 100523 (2024). 10.1016/j.xgen.2024.100523

37 Spracklen, C. N. et al. Identification of type 2 diabetes loci in 433,540 East Asian individuals. Nature 582, 240–245 (2020). 10.1038/s41586-020-2263-3

38 Chang, S. W. et al. A genome-wide association study on chronic HBV infection and its clinical progression in male Han-Taiwanese. PLoS One 9, e99724 (2014). 10.1371/journal.pone.0099724

39 Zeng, Z. et al. Genome-wide association study identifies new loci associated with risk of HBV infection and disease progression. BMC Med Genomics 14, 84 (2021). 10.1186/s12920-021-00907-0

40 Li, Y. et al. Genome-wide association study identifies 8p21.3 associated with persistent hepatitis B virus infection among Chinese. Nat Commun 7, 11664 (2016). 10.1038/ncomms11664

41 Chen, H. H. et al. Host genetic effects in pneumonia. Am J Hum Genet 108, 194–201 (2021). 10.1016/j.ajhg.2020.12.010

42 Covid-Host Genetics Initiative. A second update on mapping the human genetic architecture of COVID-19. Nature 621, E7–E26 (2023). 10.1038/s41586-023-06355-3

43 Covid-Host Genetics Initiative. A first update on mapping the human genetic architecture of COVID-19. Nature 608, E1–E10 (2022). 10.1038/s41586-022-04826-7

44 Asgari, S. & Pousaz, L. A. Human genetic variants identified that affect COVID susceptibility and severity. Nature 600, 390–391 (2021). 10.1038/d41586-021-01773-7

45 Kember, R. L. et al. Polygenic risk scores for cardiometabolic traits demonstrate importance of ancestry for predictive precision medicine. Pac Symp Biocomput 29, 611–626 (2024).

46 Khunsriraksakul, C. et al. Multi-ancestry and multi-trait genome-wide association meta-analyses inform clinical risk prediction for systemic lupus erythematosus. Nat Commun 14, 668 (2023). 10.1038/s41467-023-36306-5

47 Kelemen, M., Vigorito, E., Fachal, L., Anderson, C. A. & Wallace, C. shaPRS: Leveraging shared genetic effects across traits or ancestries improves accuracy of polygenic scores. Am J Hum Genet 111, 1006–1017 (2024). 10.1016/j.ajhg.2024.04.009

48 Zhai, S., Guo, B., Wu, B., Mehrotra, D. V. & Shen, J. Integrating multiple traits for improving polygenic risk prediction in disease and pharmacogenomics GWAS. Brief Bioinform 24 (2023). 10.1093/bib/bbad181

49 Schoeler, T. et al. Participation bias in the UK Biobank distorts genetic associations and downstream analyses. Nat Hum Behav 7, 1216–1227 (2023). 10.1038/s41562-023-01579-9

50 Chen, C.-Y. et al. Analysis across Taiwan Biobank, Biobank Japan, and UK Biobank identifies hundreds of novel loci for 36 quantitative traits. Cell Genomics 3, 100436 (2023). 10.1016/j.xgen.2023.100436

51 Wainschtein, P. et al. Assessing the contribution of rare variants to complex trait heritability from whole-genome sequence data. Nat Genet 54, 263–273 (2022). 10.1038/s41588-021-00997-7

52 Yengo, L. et al. Imprint of assortative mating on the human genome. Nat Hum Behav 2, 948–954 (2018). 10.1038/s41562-018-0476-3

53 Border, R. et al. Assortative mating biases marker-based heritability estimators. Nat Commun 13, 660 (2022). 10.1038/s41467-022-28294-9

54 Li, M. X. et al. A major gene model of adult height is suggested in Chinese. J Hum Genet 49, 148–153 (2004). 10.1007/s10038-004-0125-8

55 Zhong, Y., Perera, M. A. & Gamazon, E. R. On Using Local Ancestry to Characterize the Genetic Architecture of Human Traits: Genetic Regulation of Gene Expression in Multiethnic or Admixed Populations. Am J Hum Genet 104, 1097–1115 (2019). 10.1016/j.ajhg.2019.04.009

56 Gay, N. R. et al. Impact of admixture and ancestry on eQTL analysis and GWAS colocalization in GTEx. Genome Biol 21, 233 (2020). 10.1186/s13059-020-02113-0

57 Burch, K. S. et al. Partitioning gene-level contributions to complex-trait heritability by allele frequency identifies disease-relevant genes. Am J Hum Genet 109, 692–709 (2022). 10.1016/j.ajhg.2022.02.012

58 Wallace, C. A more accurate method for colocalisation analysis allowing for multiple causal variants. PLoS Genet 17, e1009440 (2021). 10.1371/journal.pgen.1009440

## References

59 Kirby, J. C. et al. PheKB: a catalog and workflow for creating electronic phenotype algorithms for transportability. J Am Med Inform Assoc 23, 1046–1052 (2016). 10.1093/jamia/ocv202

60 Yang, H.-C. et al. The Taiwan Precision Medicine Initiative: A Cohort for Large-Scale Studies. BIORXIV (2024). 10.1101/2024.10.14.616932v1

61 Alexander, D. H., Novembre, J. & Lange, K. Fast model-based estimation of ancestry in unrelated individuals. Genome Res 19, 1655–1664 (2009). 10.1101/gr.094052.109

62 Genomes Project Consortium et al. A global reference for human genetic variation. Nature 526, 68–74 (2015). 10.1038/nature15393

63 Hofmeister, R. J., Ribeiro, D. M., Rubinacci, S. & Delaneau, O. Accurate rare variant phasing of whole-genome and whole-exome sequencing data in the UK Biobank. Nat Genet 55, 1243–1249 (2023). 10.1038/s41588-023-01415-w

64 Rubinacci, S., Delaneau, O. & Marchini, J. Genotype imputation using the Positional Burrows Wheeler Transform. PLoS Genet 16, e1009049 (2020). 10.1371/journal.pgen.1009049

65 Conomos, M. P., Reiner, A. P., Weir, B. S. & Thornton, T. A. Model-free Estimation of Recent Genetic Relatedness. Am J Hum Genet 98, 127–148 (2016). 10.1016/j.ajhg.2015.11.022

66 Conomos, M. P., Miller, M. B. & Thornton, T. A. Robust inference of population structure for ancestry prediction and correction of stratification in the presence of relatedness. Genet Epidemiol 39, 276–293 (2015). 10.1002/gepi.21896

67 Staples, J. et al. PRIMUS: rapid reconstruction of pedigrees from genome-wide estimates of identity by descent. Am J Hum Genet 95, 553–564 (2014). 10.1016/j.ajhg.2014.10.005

68 Zhou, W. et al. Efficiently controlling for case-control imbalance and sample relatedness in large-scale genetic association studies. Nat Genet 50, 1335–1341 (2018). 10.1038/s41588-018-0184-y

69 Chang, C. C. et al. Second-generation PLINK: rising to the challenge of larger and richer datasets. Gigascience 4, 7 (2015). 10.1186/s13742-015-0047-8

70 Bastarache, L. et al. The phenotype-genotype reference map: Improving biobank data science through replication. Am J Hum Genet 110, 1522–1533 (2023). 10.1016/j.ajhg.2023.07.012

71 Zou, Y., Carbonetto, P., Wang, G. & Stephens, M. Fine-mapping from summary data with the “Sum of Single Effects” model. PLoS Genet 18, e1010299 (2022). 10.1371/journal.pgen.1010299

72 Wang, K., Li, M. & Hakonarson, H. ANNOVAR: functional annotation of genetic variants from high-throughput sequencing data. Nucleic Acids Res 38, e164 (2010). 10.1093/nar/gkq603

73 Frankish, A. et al. GENCODE: reference annotation for the human and mouse genomes in 2023. Nucleic Acids Res 51, D942–D949 (2023). 10.1093/nar/gkac1071

74 Chen, S. et al. A genomic mutational constraint map using variation in 76,156 human genomes. Nature 625, 92–100 (2024). 10.1038/s41586-023-06045-0

75 Ojavee, S. E., Kutalik, Z. & Robinson, M. R. Liability-scale heritability estimation for biobank studies of low-prevalence disease. Am J Hum Genet 109, 2009–2017 (2022). 10.1016/j.ajhg.2022.09.011

76 Lee, S. H., Goddard, M. E., Wray, N. R. & Visscher, P. M. A better coefficient of determination for genetic profile analysis. Genet Epidemiol 36, 214–224 (2012). 10.1002/gepi.21614

77 Bulik-Sullivan, B. et al. An atlas of genetic correlations across human diseases and traits. Nat Genet 47, 1236–1241 (2015). 10.1038/ng.3406

78 Wallace, C. Statistical testing of shared genetic control for potentially related traits. Genet Epidemiol 37, 802–813 (2013). 10.1002/gepi.21765

79 Wang, G., Sarkar, A., Carbonetto, P. & Stephens, M. A simple new approach to variable selection in regression, with application to genetic fine mapping. J R Stat Soc Series B Stat Methodol 82, 1273–1300 (2020). 10.1111/rssb.12388

80 Ni, G. et al. A Comparison of Ten Polygenic Score Methods for Psychiatric Disorders Applied Across Multiple Cohorts. Biol Psychiatry 90, 611–620 (2021). 10.1016/j.biopsych.2021.04.018

81 Ruan, Y. et al. Improving polygenic prediction in ancestrally diverse populations. Nature Genetics 54, 573–580 (2022). 10.1038/s41588-022-01054-7

82 Bertrand, F. & Maumy-Bertrand, M. plsRglm: Partial least squares linear and generalized linear regression for processing incomplete datasets by cross-validation and bootstrap techniques with R. 1810.01005 (2018). <https://ui.adsabs.harvard.edu/abs/2018arXiv181001005B>.

